# A systematic review of the prevalence of persistent gastrointestinal symptoms and incidence of new gastrointestinal illness after acute SARS-CoV-2 infection

**DOI:** 10.1101/2023.04.26.23289142

**Authors:** Michael J. Hawkings, N. Marcella Vaselli, Dimitrios Charalampopoulos, Liam Brierley, Alex J. Elliot, Iain Buchan, Daniel Hungerford

**Affiliations:** Institute of Population Health, Department of Public Health, Policy & Systems, University of Liverpool, Liverpool, L69 3GF, UK; National Institute for Health and Care Research Health Protection Research Unit in Gastrointestinal Infections at University of Liverpool, Liverpool, L69 7BE, UK; Institute of Infection, Veterinary & Ecological Sciences, Department of Clinical Infection, Microbiology and Immunology, University of Liverpool, Liverpool, L69 7BE, UK; Institute of Population Health, Department of Health Data Science, University of Liverpool, Liverpool, L69 3GF, UK; Real-Time Syndromic Surveillance Team, Field Services, Health Protection Operations, UK Health Security Agency, Birmingham, B2 4BH, UK

**Keywords:** SARS-CoV-2, long-COVID, post-COVID syndrome, gastrointestinal, diarrhoea, irritable bowel syndrome

## Abstract

It is known that SARS-CoV-2 infection can result in gastrointestinal symptoms. For some, these symptoms may persist beyond acute infection, in what is known as ‘post-COVID syndrome’. We conducted a systematic review to examine the prevalence of persistent gastrointestinal symptoms and the incidence of new gastrointestinal illness following acute SARS-CoV-2 infection. We searched scientific literature using MedLine, SCOPUS, Embase, Europe PubMed Central, medRxiv and Google Scholar from December 2019 to October 2022. Two reviewers independently identified 28 eligible articles which followed participants for various gastrointestinal outcomes after acute SARS-CoV-2 infection. Study quality was assessed using the Joanna Briggs Institute Critical Appraisal Tools. The weighted pooled prevalence for persistent gastrointestinal symptom of any nature and duration was 10.7%, compared to 4.9% in healthy controls. For six studies at a low risk of methodological bias, the symptom prevalence ranged from 0.2% to 24.1% with a median follow-up time of 13 weeks. We also identified the presence of functional gastrointestinal disorders in historically SARS-CoV-2 exposed individuals. Our review has shown that, from a limited pool of mostly low-quality studies, previous SARS-CoV-2 exposure may be associated with ongoing gastrointestinal symptoms and the development of functional gastrointestinal illness. Furthermore, we show the need for high-quality research to better understand the SARS-CoV-2 association with gastrointestinal symptoms, particularly as population exposure to enteric infections returns to pre-COVID-19-restriction levels.

**Highlights:**

- Acute SARS-CoV-2 infection can result in gastrointestinal symptoms
- The burden of gastrointestinal illness after acute SARS-CoV-2 infection is not known
- This systematic review evaluates the evidence across 28 observational studies
- Most studies identified are at high risk of bias and of low quality
- SARS-CoV-2 exposure may be associated with new post-infection gastrointestinal illness

## Introduction

Since its emergence in late 2019, the COVID-19 pandemic has resulted in over 650 million infections and 6 million deaths worldwide as of January 2023^1^. While most cases are experienced as a self-limiting respiratory tract infection, some individuals may develop a more severe or protracted illness. Post-COVID-19 syndrome, or long-COVID, refers to symptoms that persist beyond 12 weeks from the onset of infection^2^. In the UK, self-reported health data indicate that 2.1 million people, or 3.3% of the general population, were suffering from post-COVID in January 2023^3^. Symptoms of post-COVID syndrome appear to affect every body system, often overlapping, and with a significant mental health impact^2^.

It is established that SARS-CoV-2 (the virus that causes COVID-19 disease) infection can result in gastrointestinal symptoms: in 2020, a meta-analysis of 8302 patients identified diarrhoea in 12% of paediatric and 9% of adult cases^4^. Elshazli et al. also identified gastrointestinal symptoms in 20% of 25,252 patients, with anorexia, dysgeusia, diarrhoea, nausea and haematemesis being most common^5^. In Liverpool, UK, gastrointestinal symptoms were observed in a third of hospitalised COVID-19 patients^6^. Research investigating gastrointestinal symptoms as a component of post-COVID syndrome, however, is less substantial. One review of post-COVID syndrome patients identified persistent symptoms of dysgeusia and diarrhoea at a frequency of 7% and 6% respectively^7^. Similarly, an electronic health record study of 273,618 individuals in the USA found persistent gastrointestinal symptoms in 8.3% of participants six months after COVID-19 onset^8^. Chopra et al. also found that patients presenting with diarrhoea during acute COVID-19 were more likely to suffer from post-COVID syndrome symptoms such as fatigue, dyspnoea and chest discomfort^9^.

When considering how SARS-CoV-2 interacts with the gastrointestinal system, and how this may result in persistent post-viral symptoms, several pathophysiological mechanisms have been proposed. SARS-CoV-2 initially binds with the angiotensin converting enzyme-2 (ACE-2) receptor, which is highly expressed by ileal enterocytes^10, 11^. The binding of SARS-CoV-2 to ACE-2 receptors may disrupt angiotensin homeostasis and reduce tryptophan absorption, resulting in inflammation and alteration of the gut microbiota^12, 13^. Alteration of the gut microbiome is known to occur during acute COVID-19 infection, as it does in many other viral respiratory tract infections^14, 15^. This can result in the depletion of beneficial gut commensals and the proliferation of opportunistic pathogens in the gastrointestinal tract^16^. Zuo et al. found this dysbiotic state to worsen over time, even after recovery from the acute illness, in moderate to severe hospitalised COVID-19 cases^17^. Phetsouphanh et al. identified raised levels of IFN-β, IFN-λ1 and highly activated innate immune cells eight months after COVID-19 diagnosis, while other studies detected the persistence of SARS-CoV-2 viral matter in the intestinal epithelium beyond recovery^18–20^. Another mechanism that has been recently suggested is the formation of fibrinolysis-resistant amyloid microclots and platelet pathology in post-COVID syndrome patients^21^. Kell et al. proposed that these microclots may impair tissue perfusion and be a key determinant of post-COVID syndrome^22^.

Although, to the best of the authors’ knowledge, no studies thus far have investigated the mental wellbeing impact of persistent gastrointestinal post-COVID symptoms, the psychosocial impact of functional gastrointestinal disorders (FGIDs) is known to be significant. Similarity between FGIDs and post-COVID gastrointestinal symptoms can be drawn in their clinical features and poorly understood causal mechanisms, both likely including the intestinal microbiota and gut-brain axis^23^. Around half of irritable bowel syndrome (IBS) patients are thought to have concurrent mental health conditions, and symptom-specific anxiety can impair functions of daily life in affected patients^24^. The development of low mood and anxiety states has also been observed in post-COVID syndrome^25^. Given the potential for a substantial physical and mental disease burden of both post-COVID syndrome and gastrointestinal illness, we conducted a systematic review aiming to summarising the current evidence in two areas. Firstly, to provide an overview of the prevalence of persistent gastrointestinal symptoms following acute SARS-CoV-2 infection, and secondly, to estimate the incidence of newly diagnosed gastrointestinal illness following recent SARS-CoV-2 infection (not including gastrointestinal complications of acute COVID-19).

## Methods

### Search Strategy

The review was conducted in accordance with the Preferred Reporting Items for Systematic Reviews and Meta-Analyses (PRISMA) standards for systematic reviews, and registered with the International Register of Systematic Reviews (PROSPERO reference CRD42022315792, available from: https://www.crd.york.ac.uk/prospero/display_record.php?ID=CRD42022315792).

We conducted a systematic search of the literature using OVID MedLine, SCOPUS and Europe PubMed Central from 01 December 2019 to 11 October 2022. We also searched pre-publication and other literature indexed in medRxiv and Google Scholar. Search terms were constructed around three themes: COVID-19, gastrointestinal symptoms or illness and observational study designs. We did not include terms such as ‘obesity’ due to the volume of unsuitable studies returned. Our search terms were reviewed by all authors and a health science librarian. Our full search strategy is shown in table 1.

**Table 1.** Overview of search strategy including limits.

| Search Strategy |  |
| --- | --- |
| 1 | "COVID-19" [Mesh] OR "SARS CoV-2" OR "coronavirus disease 2019" |
| 2 | "Gastrointestinal Disease" [Mesh] OR "gastrointestinal symptoms" OR "abdominal pain" OR "nausea" OR "vomiting" OR "diarrhea" OR "Diarrhea, Infantile" [Mesh] OR "constipation" OR "malnutrition" OR "gastritis" OR "gastroesophageal reflux" OR "pancreatitis" OR "colitis" OR "Deglutition Disorders" [Mesh] OR "esophagitis" OR "transaminitis" OR "cholestasis" OR "cholestatic liver injury" OR "appendicitis" OR "Gastrointestinal Haemorrhage" [Mesh] OR "upper gastrointestinal bleed" OR "lower gastrointestinal bleed" |
| 3 | "Longitudinal Studies" [Mesh] OR "longitudinal" OR "Cross-Sectional Studies" [Mesh] OR "cross-sectional" OR "Cohort Studies" OR "cohort" OR "Case-Control Studies" [Mesh] OR "case control" OR "Observational Studies" [Mesh] OR "observational" |
| 4 | 1 AND 2 AND 3 |
| 5 | Limit to humans and English language |
| 6 | 4 AND 5 |

### Inclusion/Exclusion Criteria

Studies were included in the review if they followed up participants for ongoing gastrointestinal symptoms or new gastrointestinal illness beyond acute COVID-19 infection. We used the following inclusion criteria: (1) follow-up of gastrointestinal symptoms and the development of new gastrointestinal illness from 4 weeks after COVID-19 diagnosis or onset, as per the National Institute for Health and Care Excellence (NICE) UK case definition for acute COVID-19 infection^2^; (2) observational studies, including cohort, case-control and cross sectional studies; and (3) published in English. We excluded studies that: (1) were conducted before December 2019; (2) were case reports, opinions, commentaries and interventional studies; (3) included unconfirmed COVID-19 or other SARS-like illnesses; (4) involved animals; and (5) did not meet the inclusion criteria. Pre-print articles were considered acceptable if they met the inclusion criteria.

We anticipated studies reporting persistent symptom prevalence would involve participants reporting a gastrointestinal symptom during acute infection and its persistence beyond four weeks from diagnosis, or beyond the resolution of other COVID-19 symptoms. While symptom persistence beyond twelve weeks may warrant a diagnosis of post-COVID syndrome, this diagnostic term was not used earlier in the pandemic and would not detect participants with post-viral symptoms lasting for between four and twelve weeks. Due to nature of review topic, we imposed no restrictions on participant age or comorbidity, and studies were not required to include a control or comparator group for inclusion. Two reviewers (MJH and NMV) independently screened the citations, and any discrepancies were resolved by consulting a third reviewer (DH).

### Data Extraction

The following data were extracted into a spreadsheet for manual review:

- Study details: publication date, journal, authors, year, location, study design and setting (i.e. community or hospital), funding source.
- Population characteristics: age, number of cases and controls, case definition, illness severity, vaccination status, SARS-CoV-2 variant, diagnostic criteria.
- Acute COVID-19 symptoms relating to the gastrointestinal system.
- Point prevalence of persistent gastrointestinal symptoms after acute COVID-19, and the timepoint and method for which these symptoms were reported. Post-COVID symptoms reported would be persistent in nature i.e. participants with a short-lived, unrelated episode of acute gastroenteritis at follow-up would not be captured.
- Incidence of new gastrointestinal illness presenting after recovery from acute COVID-19.

### Risk of Bias and Quality Assessment

Studies eligible for inclusion were quality assessed using the Joanna Briggs Institute (JBI) Critical Appraisal Tools for observational studies^26^. Quality and risk of bias was assessed by two reviewers independently (MJH and NMV). The JBI Critical Appraisal Tools assesses the methodological quality and risk of bias of each study, appraising aspects such as exposure and outcome measurement, controlling for confounding variables, loss of participants to follow-up and appropriateness of statistical analysis. Strength of methodological quality was graded by the percentage of positive answers, with studies scoring ≥70% considered high quality, studies scoring 50-69% considered moderate quality and studies scoring ≤49% considered low quality^27, 28^.

### Data Analysis

We conducted a narrative synthesis of the studies to summarise the characteristics of each study, method of diagnosis and gastrointestinal-specific outcome measurements. We then conducted a descriptive analysis of all studies, and reported the point prevalence for each symptom and the time at which this was captured. Where possible, persistent symptoms were grouped into one of four categories: diarrhoea (including loose stools and liquid stools); nausea and vomiting (including feeling and being sick); taste and smell disorders (including ageusia, dysgeusia, anosmia and altered taste and smell); and abdominal pain (including stomach ache/pain). We estimated the pooled prevalence for each symptom category across all studies by calculating the weighted average of the number of symptomatic cases divided by the total cohort size.

Change in point prevalence was visualised against time from acute COVID-19 resolution. Where participants reported more than one symptom per category (i.e. nausea, vomiting or haematemesis), the highest reported prevalence was used in the analysis. Overall pooled prevalence for any gastrointestinal complaint was only calculated from studies which reported this specifically, and not calculated from the raw data to avoid double counting of participants reporting multiple symptoms.

For studies reporting the prevalence of gastrointestinal symptoms in both COVID-19 cases and controls, we calculated the odds ratio of having persistent gastrointestinal symptoms in a COVID-19 cohort *vs.* healthy controls for each symptom in each study. We calculated the I^2^ statistic to assess heterogeneity between studies and conducted both common and random effects meta-analyses to estimate overall odds ratios and 95% confidence intervals using the ‘metabin’ function in R package ‘meta’, version 6.0-0 in R version 4.1.0^29^.

## Results

The initial search after duplicate removal identified 1,488 potentially relevant studies; of these, 28 were eligible for inclusion in the review. Each reported the point prevalence of persistent gastrointestinal symptoms at specific timepoints, up to a maximum of 12 months from SARS-CoV-2 infection (Figure 1). The studies were conducted from early 2020 to mid-2022 in fifteen different countries across Europe, North and Central America, China and India (Table 2 and 3). Twenty-six studies were sourced from peer-review journals^8, 30–54^. Two studies were pre-print and were awaiting peer review at the time of writing^55, 56^.

**Fig 1.**
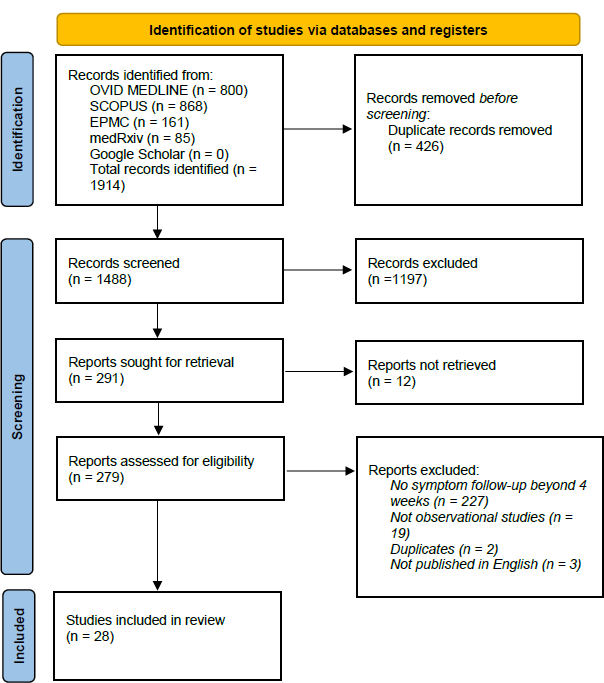
PRISMA flowchart detailing search results.

**Table 2.**
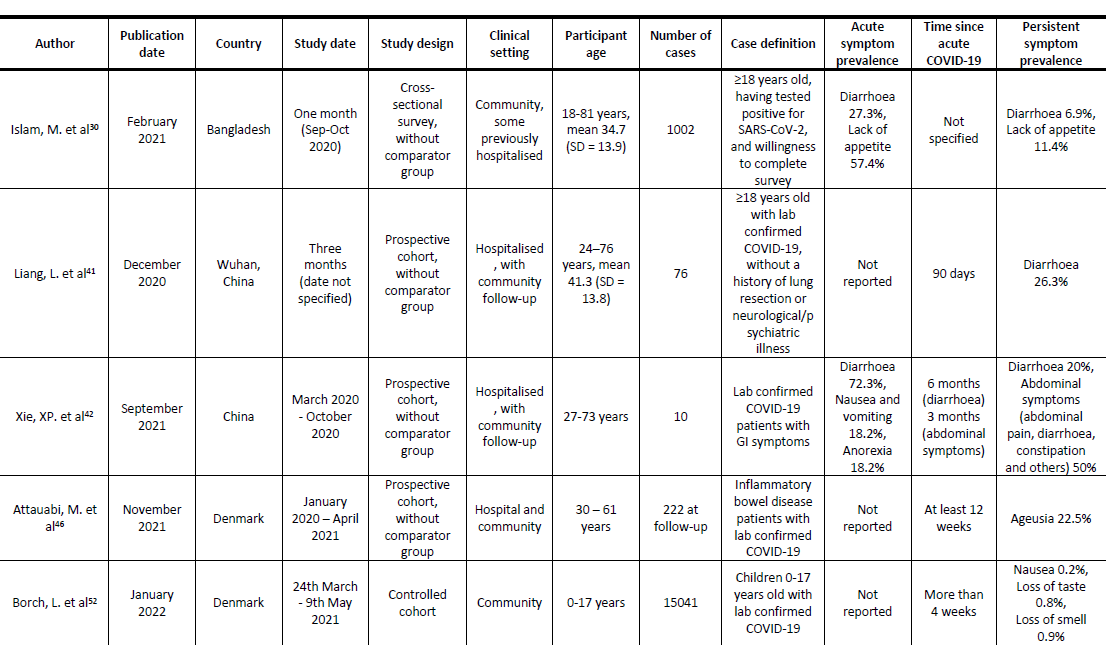
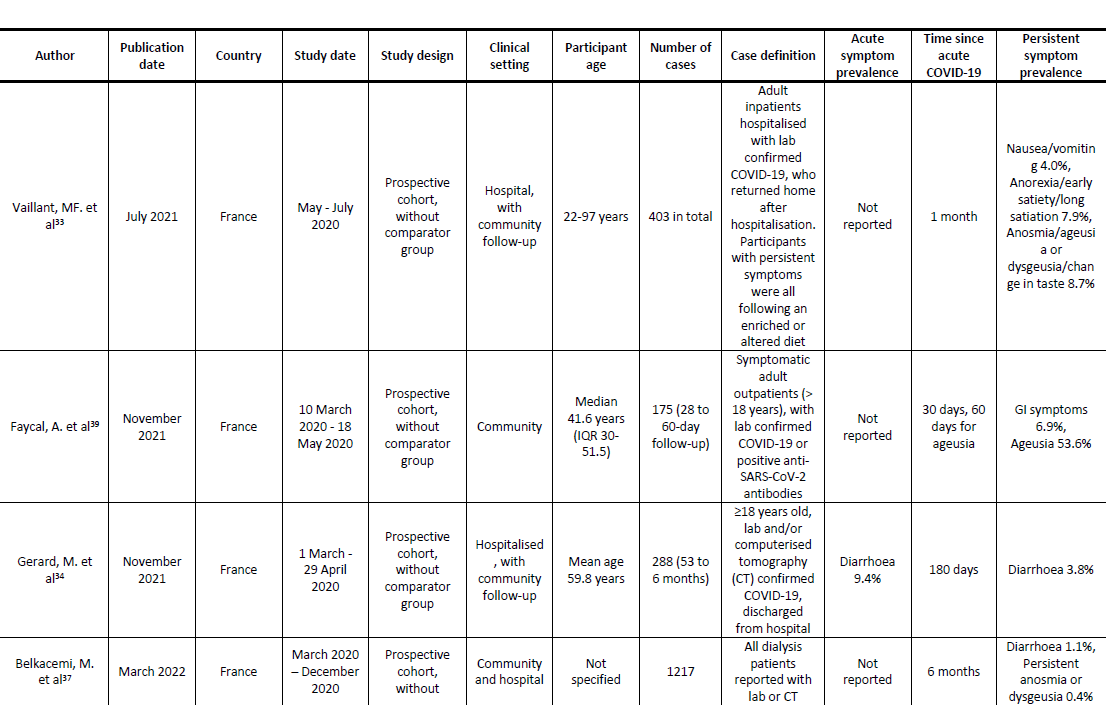
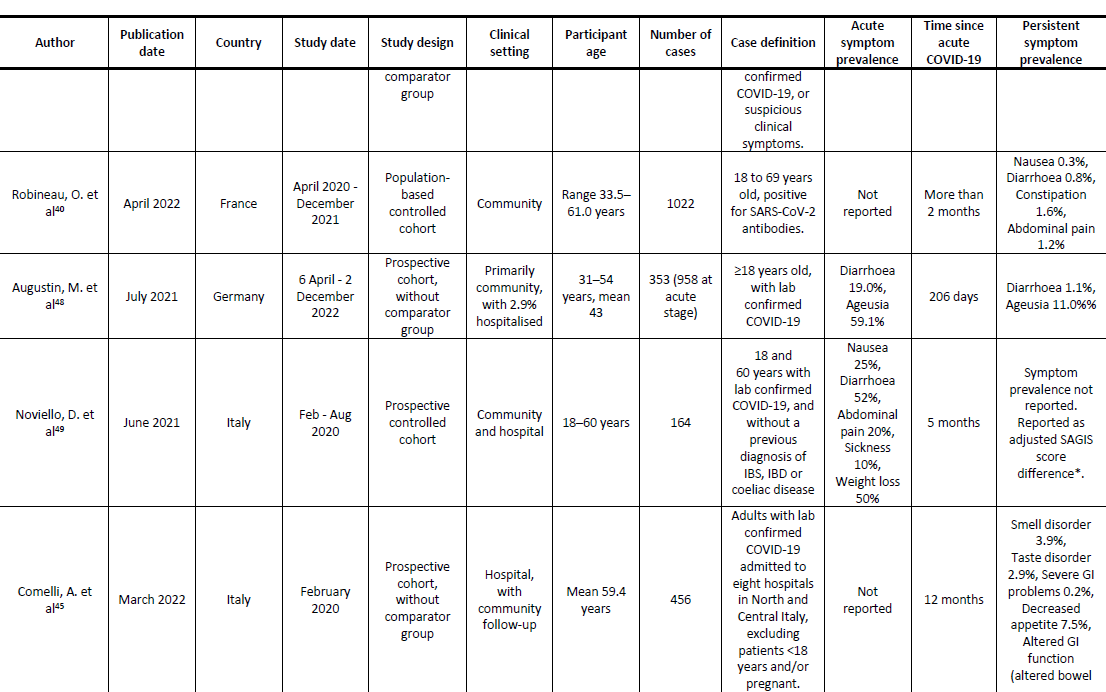
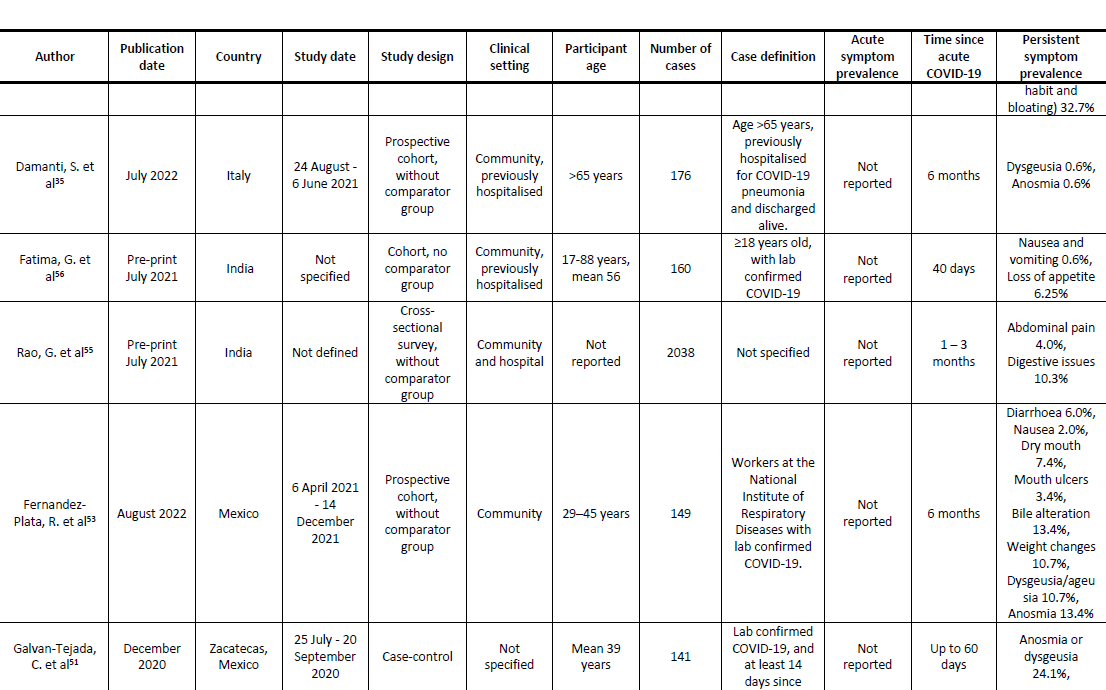
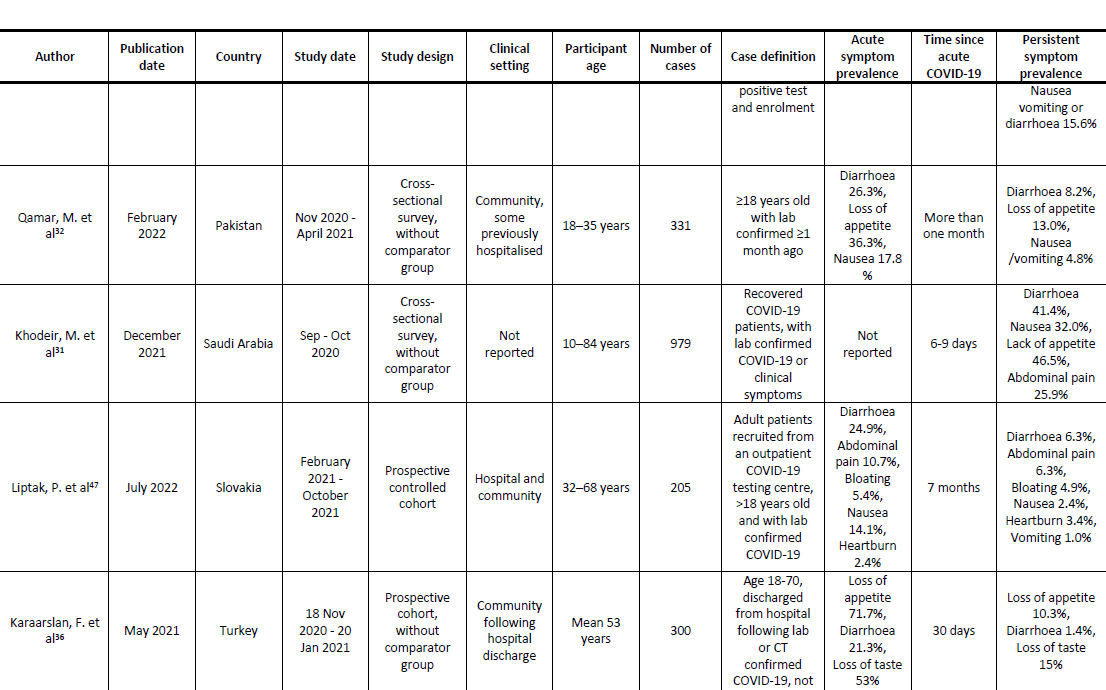
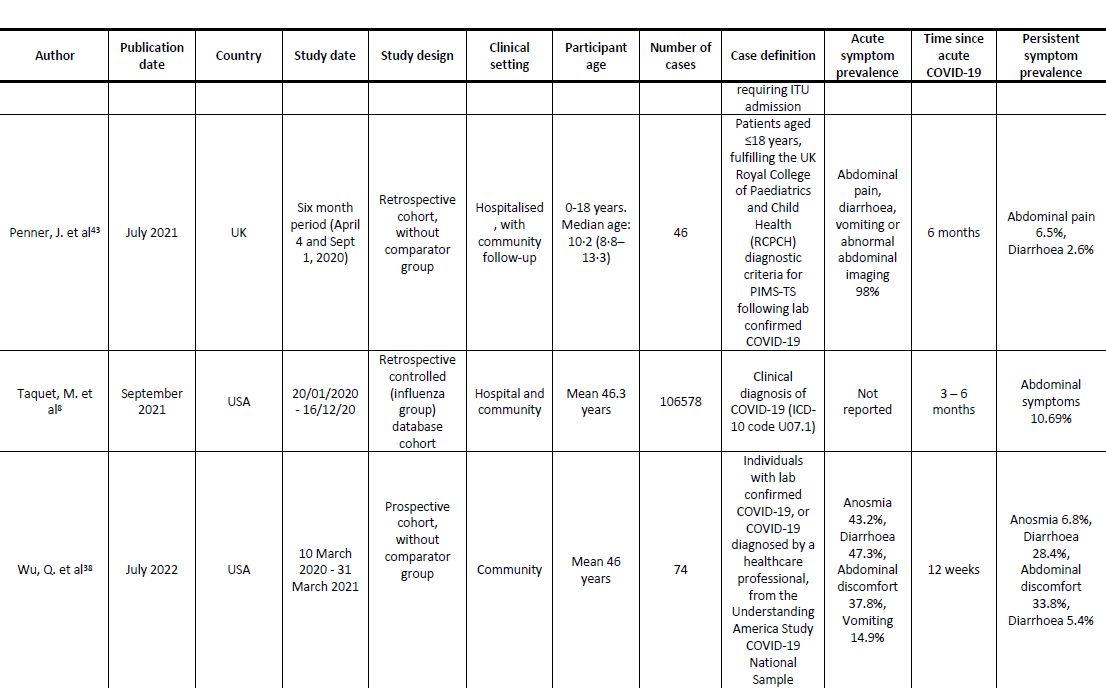
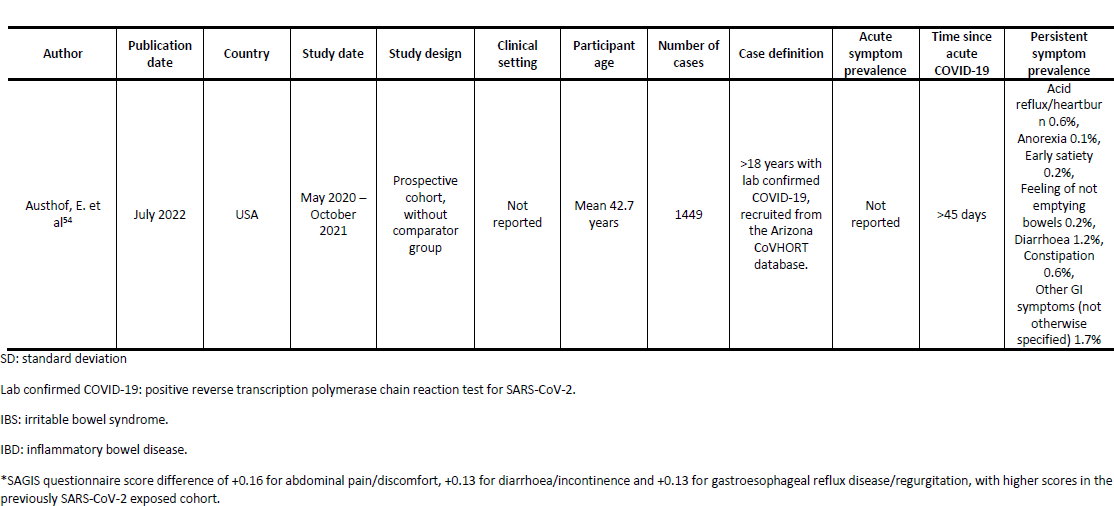
Descriptive sucmmary of studies reporting the point prevalence of persistent gastrointestinal symptoms after acute COVID-19.

**Table 3.**
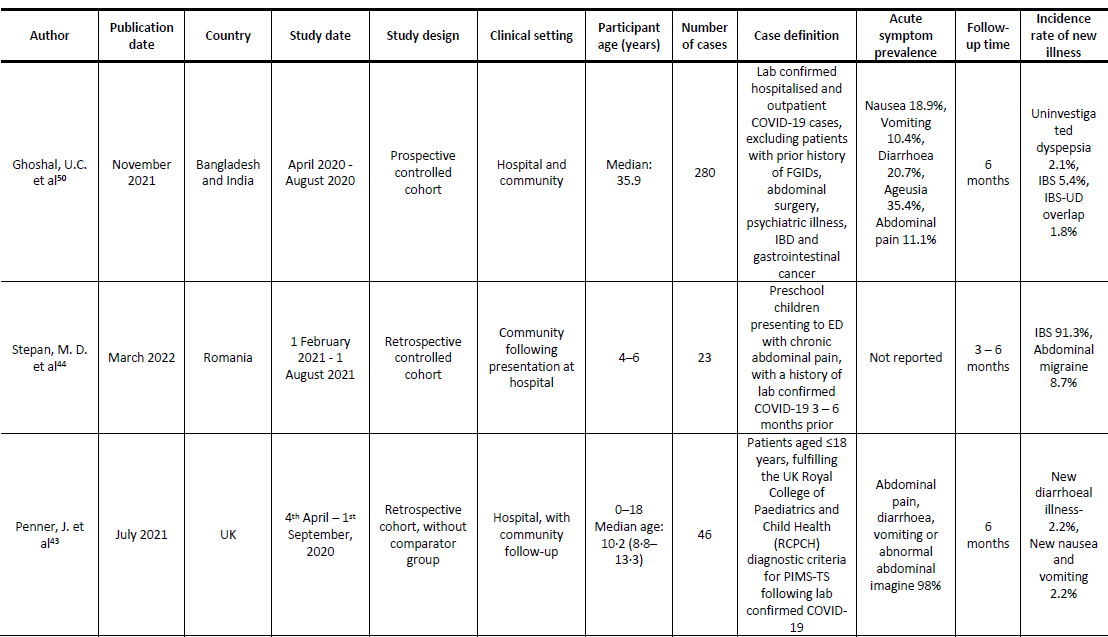
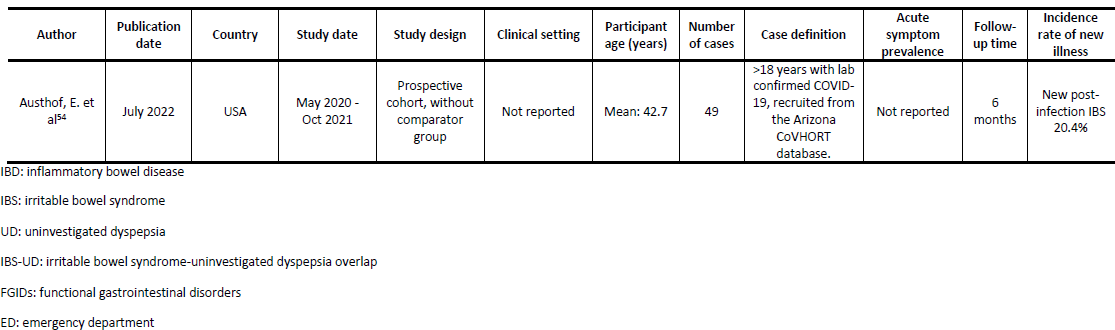
Descriptive summary of studies reporting the incidence of new gastrointestinal illness after acute SARS-CoV-2 infection.

### Study characteristics and outcome measurement

Most studies (n = 20) followed up participants from COVID-19 diagnosis or acute illness to recovery (Table 2 and 3) and assessed for post-viral symptoms thereafter. Four studies followed up patients for a different clinical reason, such as nutritional status and endoscopy, and assessed post-COVID symptoms as a secondary outcome. The remaining four studies were cross-sectional and reported the point prevalence of a persistent symptom in participants who had historically tested positive for SARS-CoV-2^30–32, 55^. Three of these studies had a maximum possible duration between diagnosis and inclusion in the study of 16 months, although for most participants this was less than six months^30–32^. One cross-sectional study did not specify the study period or time to follow-up^55^.

Studies varied in how they defined COVID-19 cases. Sixteen studies included only participants with a positive reverse transcription polymerase chain reaction test (RT-PCR) for SARS-CoV-2^32, 41–54, 56^. Three studies included participants who tested positive via an unspecified diagnostic method^30, 33, 55^. One retrospective database cohort study included participants with an ICD-10 code for confirmed COVID-19 in their electronic health record (EHR)^8^. Three studies included patients with COVID-19 pneumonia diagnosed through computerised tomography (CT) scan in addition to those diagnosed via RT-PCR^34–36^. Two included those diagnosed on clinical symptoms in addition to those diagnosed via RT-PCR and CT scan, but this formed a small percentage of the overall cohort (4% and 16%)^31, 37^. One study included participants with COVID-19 illness diagnosed based on clinical history and examination by a physician, in addition to those with laboratory confirmed SARS-CoV-2 infection^38^. Two studies included participants with positive anti-SARS-CoV-2 antibodies on serological testing, with one also including participants diagnosed via RT-PCR^39, 40^. No studies reported SARS-CoV-2 vaccination status, however, it is likely the vast majority of participants were not vaccinated due to the timing of studies in relation to vaccine rollout globally. The studies also varied in their clinical settings and ascertainment of gastrointestinal symptoms. Nine studies were conducted in patients who were hospitalised with COVID-19, discharged, and followed-up in the community^33–35, 41–45, 56^, twelve studies involved both hospitalised and community-managed cases^8, 30, 32, 36, 37, 40, 46–50, 55^, five studies involved community cases that did not require hospital admission^38, 39, 51–53^ and two studies did not specify the setting of recruitment^31, 54^.

Most studies (n = 19) measured gastrointestinal outcomes using a survey or questionnaire, administered via telephone or electronically^30–32, 36–40, 45–47, 49–56^. One study used EHRs to identify gastrointestinal endpoints based on clinical codes^8^. Six studies involved an in-person clinical assessment by a healthcare professional^33–35, 41, 43, 44^. One study involved both a questionnaire and clinical assessment with a healthcare professional^48^. One study measured outcomes through both clinical assessment and endoscopy^42^.

### Symptom prevalence

Overall, the average prevalence of persistent post-COVID gastrointestinal symptoms of any nature and duration was 28.8%, or 10.7% when weighted by cohort size (n = 110,886 across six studies). For two studies reporting the total prevalence of persistent gastrointestinal symptoms of any nature and duration in healthy controls, weighted average prevalence was 4.9% (n = 106,710).

Figure 2 shows the prevalence of persistent gastrointestinal symptoms over time in adults. We distinguished between studies that report point prevalence at a specific time point and studies that reported prevalence with minimum symptom duration (i.e. diarrhoea at 12 weeks *vs.* diarrhoea for at least 12 weeks). For studies reporting symptom prevalence at multiple timepoints, we selected the latest timepoint to avoid double counting of participants. We calculated pooled symptom prevalence for studies reporting diarrhoea, nausea and vomiting, taste and smell disorders and abdominal pain, persisting for between one and 12 months (Table 4).

**Figure 2.**
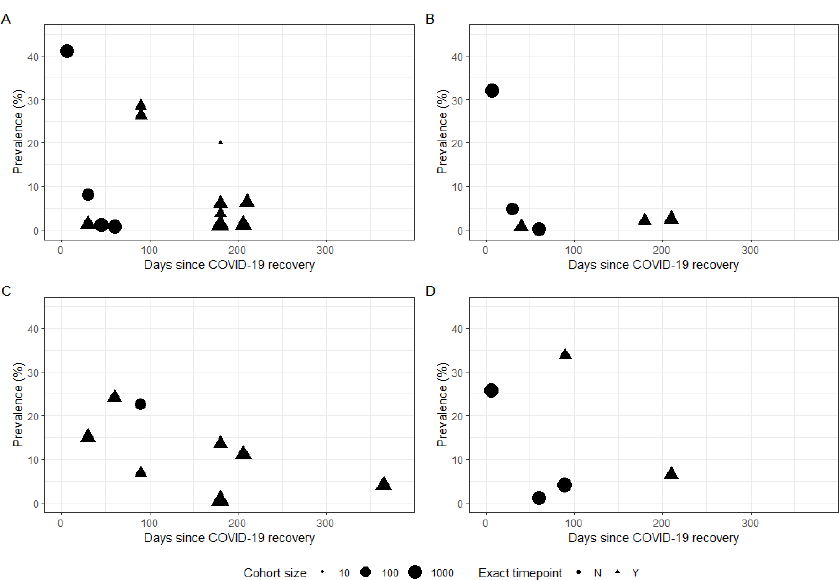
Time plots showing symptom prevalence against time since acute SARS-CoV-2 infection in adults. (A): prevalence of diarrhoea. (B): prevalence of nausea and vomiting. (C): prevalence of taste and smell disorders. (D): prevalence of abdominal pain. Size of points indicates cohort size. Triangles indicate point prevalence at that exact timepoint, whereas circles indicate studies reporting the lower bound of symptom duration.

**Table 4.** Average pooled prevalence of persistent symptoms for any duration between 1 month to 12 months.

| Symptom | Total sample size | Weighted prevalence | Unweighted prevalence |
| --- | --- | --- | --- |
| Diarrhoea | 22,308 across 16 studies | 2.8% | 9.6% |
| Nausea and vomiting | 18,035 across 8 studies | 2.1% | 6.6% |
| Taste and smell disorders | 18,305 across 12 studies | 2.2% | 15.0% |
| Abdominal pain | 4364 across 6 studies | 8.9% | 12.9% |

Four studies reported the prevalence of diarrhoea, nausea and vomiting, taste and smell disorders and abdominal pain in both a SARS-CoV-2 exposed cohort and an unexposed control group. We could not calculate a meta-analysis of odds ratios reliably for these studies due to the low number of studies including a comparator group and significant inter-study heterogeneity (I^2^ = 79 – 83% for each symptom).

### Studies with the highest quality score

Four studies in adults scored very highly for methodological quality and were deemed to be at a low risk of bias (Table 5). The majority of studies scored poorly due to the absence of a comparator group. Noviello et al. compared persistent gastrointestinal symptoms between a cohort five months after acute COVID-19 and a cohort of healthy controls using the Structured Assessment of Gastrointestinal Symptoms (SAGIS) questionnaire, a validated tool used to identify the presence of a broad range of gastrointestinal illnesses^49, 57^. There was a higher prevalence of diarrhoea in the COVID-19 cohort at five months than in healthy controls (21.2% *vs* 9.6%, *p* = 0.05). While the incidence of IBS was similar in both cohorts (adjusted risk ratio 1.07 [95% CI: 0.72 – 1.60]), the prevalence of diarrhoea was higher in the COVID-19 cohort (adjusted risk ratio 1.88 [95% CI: 0.99 – 3.54])^49^.

**Table 5.**
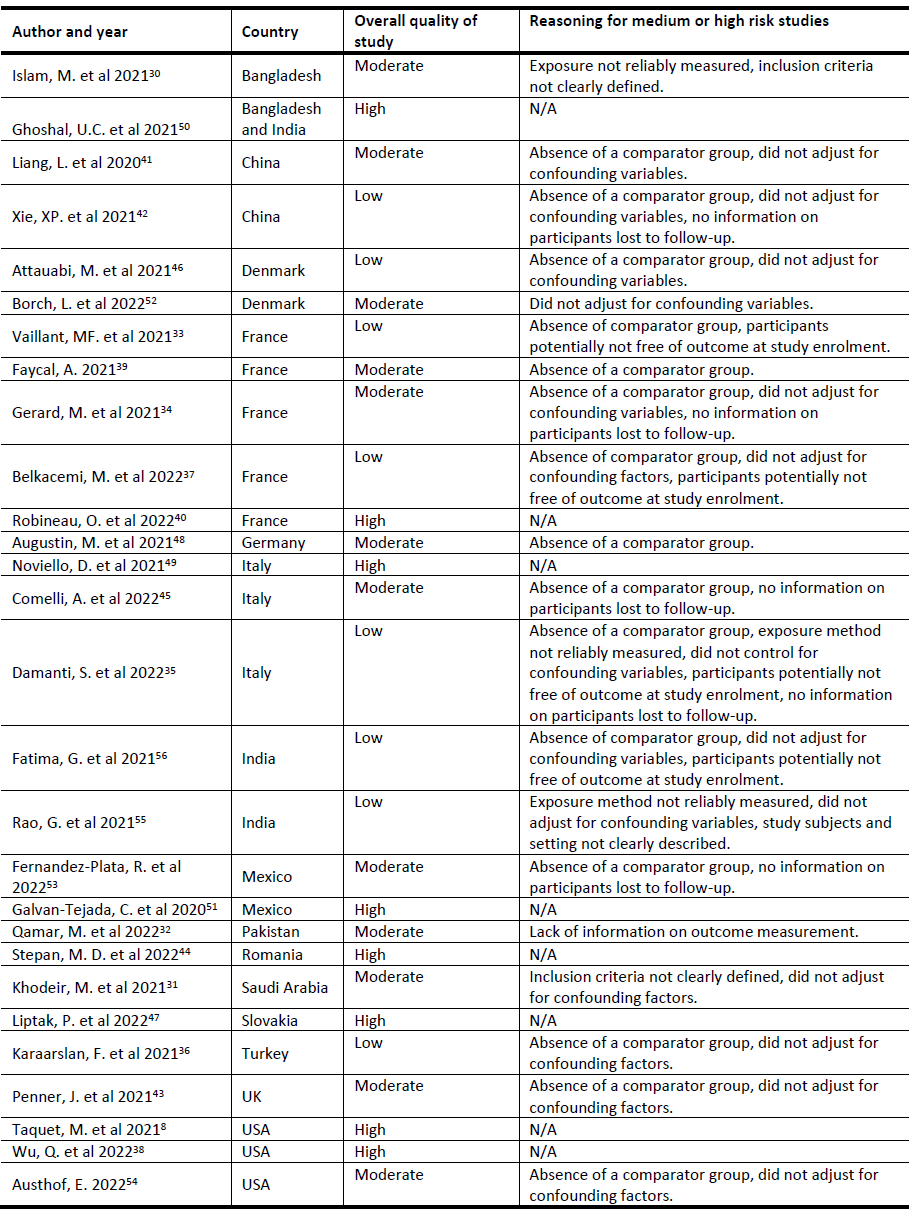
Quality and risk of bias assessment for included studies.<colcnt=4>

| Author and year | Country | Overall quality of study | Reasoning for medium or high risk studies |
| --- | --- | --- | --- |
| Islam, M. et al 2021<sup>30</sup> | Bangladesh | Moderate | Exposure not reliably measured, inclusion criteria not clearly defined. |
| Ghoshal, U.C. et al 2021<sup>50</sup> | Bangladesh and India | High | N/A |
| Liang, L. et al 2020<sup>41</sup> | China | Moderate | Absence of a comparator group, did not adjust for confounding variables. |
| Xie, XP. et al 2021<sup>42</sup> | China | Low | Absence of a comparator group, did not adjust for confounding variables, no information on participants lost to follow-up. |
| Attauabi, M. et al 2021<sup>46</sup> | Denmark | Low | Absence of a comparator group, did not adjust for confounding variables. |
| Borch, L. et al 2022<sup>52</sup> | Denmark | Moderate | Did not adjust for confounding variables. |
| Vaillant, MF. et al 2021<sup>33</sup> | France | Low | Absence of comparator group, participants potentially not free of outcome at study enrolment. |
| Faycal, A. 2021<sup>39</sup> | France | Moderate | Absence of a comparator group. |
| Gerard, M. et al 2021<sup>34</sup> | France | Moderate | Absence of a comparator group, did not adjust for confounding variables, no information on participants lost to follow-up. |
| Belkacemi, M. et al 2022<sup>37</sup> | France | Low | Absence of comparator group, did not adjust for confounding factors, participants potentially not free of outcome at study enrolment. |
| Robineau, O. et al 2022<sup>40</sup> | France | High | N/A |
| Augustin, M. et al 2021<sup>48</sup> | Germany | Moderate | Absence of a comparator group. |
| Noviello, D. et al 2021<sup>49</sup> | Italy | High | N/A |
| Comelli, A. et al 2022<sup>45</sup> | Italy | Moderate | Absence of a comparator group, no information on participants lost to follow-up. |
| Damanti, S. et al 2022<sup>35</sup> | Italy | Low | Absence of a comparator group, exposure method not reliably measured, did not control for confounding variables, participants potentially not free of outcome at study enrolment, no information on participants lost to follow-up. |
| Fatima, G. et al 2021<sup>56</sup> | India | Low | Absence of comparator group, did not adjust for confounding variables, participants potentially not free of outcome at study enrolment. |
| Rao, G. et al 2021<sup>55</sup> | India | Low | Exposure method not reliably measured, did not adjust for confounding variables, study subjects and setting not clearly described. |
| Fernandez-Plata, R. et al 2022<sup>53</sup> | Mexico | Moderate | Absence of a comparator group, no information on participants lost to follow-up. |
| Galvan-Tejada, C. et al 2020<sup>51</sup> | Mexico | High | N/A |
| Qamar, M. et al 2022<sup>32</sup> | Pakistan | Moderate | Lack of information on outcome measurement. |
| Stepan, M. D. et al 2022<sup>44</sup> | Romania | High | N/A |
| Khodeir, M. et al 2021<sup>31</sup> | Saudi Arabia | Moderate | Inclusion criteria not clearly defined, did not adjust for confounding factors. |
| Liptak, P. et al 2022<sup>47</sup> | Slovakia | High | N/A |
| Karaarslan, F. et al 2021<sup>36</sup> | Turkey | Low | Absence of a comparator group, did not adjust for confounding factors. |
| Penner, J. et al 2021<sup>43</sup> | UK | Moderate | Absence of a comparator group, did not adjust for confounding factors. |
| Taquet, M. et al 2021<sup>8</sup> | USA | High | N/A |
| Wu, Q. et al 2022<sup>38</sup> | USA | High | N/A |
| Austhof, E. 2022<sup>54</sup> | USA | Moderate | Absence of a comparator group, did not adjust for confounding factors. |

Liptak et al. followed up patients with moderate to severe COVID-19 and patients with mild COVID-19 seven months after infection and compared them with test-negative controls who visited a hospital emergency department. They reported at least one gastrointestinal symptom in 19% of the moderate to severe group and 7.3% of the mild group, while only 3.0% of the controls had any gastrointestinal symptom (*p* = <0.01). Diarrhoea and abdominal pain were more prevalent in the COVID-19 cohorts *vs.* test negative controls (*p* <0.05 for diarrhoea and *p* <0.001 for abdominal pain)^47^.

Robineau et al. assessed for post-COVID syndrome symptoms using a questionnaire in 25,910 participants after performing home dried blood spot testing for anti-SARS-CoV-2 antibodies. The presence of nausea, diarrhoea and constipation was weakly associated with SARS-CoV-2 seronegativity (adjusted odds ratios: 0.68 [95% CI: 0.16 – 1.95] for nausea; 0.61 [95% CI: 0.26 – 1.27] for diarrhoea; and 0.78 [95% CI: 0.42 – 1.33] for constipation). Abdominal pain lasting more than two months was significantly associated with negative SARS-CoV-2 serology (adjusted OR 0.42 [95% CI: 0.21 – 0.74], *p* = 0.006). Persistent anosmia or dysgeusia was strongly associated with seropositivity (adjusted OR 8.89 [95% CI: 6.03 – 13.28], *p* <0.0001)^40^.

### Studies conducted in specific patient groups

Two studies were conducted in patient cohorts. One study reported the prevalence of post-COVID symptoms in a cohort of 222 inflammatory bowel disease patients and found that 42.3% of ulcerative colitis (UC) and 45.9% of Crohn’s disease (CD) had symptoms lasting for more than twelve weeks. The most common persistent symptoms were abdominal pain (∼11% in CD and ∼4% in UC), diarrhoea (∼8% in CD and ∼3% in UC), and nausea (∼7% in CD and ∼3% in UC). Vomiting was less common (∼0-1% in both groups). Symptoms persistence beyond 12 weeks was associated with discontinuation of immunosuppressive therapy in UC patients and initial hospitalisation in CD patients^46^.

Belkacemi et al. investigated persistent post-COVID symptoms in 1217 unvaccinated end-stage renal disease patients undergoing renal replacement therapy, and reported the point prevalence of diarrhoea and taste and smell disorders at six months as 6% and 2.3% respectively (n = 216). The risk of having persisting clinical symptoms at six months was higher in patients who were hospitalised with moderate to severe disease (1.64 times) and those requiring intensive care treatment (5.03 times). Older age and longer duration of dialysis was also found to increase the risk of persistent symptoms^37^.

### Children

Two studies included children under 18 years old. Borch et al reported that, in children aged 6-17 years, diarrhoea and nausea were significantly less prevalent in the SARS-CoV-2 exposed group than in the control group (OR: 0.37, 95% CI: 0.18-0.73) one month after acute COVID-19 recovery^52^. Penner at al. found that 6.5% and 2.6% of children who has been hospitalised for paediatric multisystem inflammatory syndrome (n = 46) experienced abdominal pain and diarrhoea, respectively, at six months. This study also reported raised calprotectin, a biomarker found in faeces indicating gut inflammation, in 31% of children at six weeks, and 7% at six months^43^.

### Incidence of gastrointestinal illness

Four studies report the incidence of new gastrointestinal disease following previous SARS-CoV-2 infection^43, 44, 50, 54^. No studies reported the incidence of gastrointestinal infection following acute COVID-19. Stepan et al. found that children who visited the emergency department with abdominal pain, and tested positive for SARS-CoV-2 either six months before or three months after, had a higher incidence of irritable bowel syndrome (IBS) than those who tested negative (91.3% *vs.* 54.5%, *p* = 0.044)^44^.

Ghoshal et al. reported the incidence of new post-infection IBS, uninvestigated dyspepsia (UD) and IBS-UD overlap to be 5.3%, 2.1% and 1.8% respectively (n = 280), in those who tested positive for SARS-CoV-2 six months prior. The incidence of IBS in the control group was 0.4%, and no controls were diagnosed with UD (n = 264). Kaplan-Meier analysis showed a higher probability of developing such illnesses following SARS-CoV-2 exposure *vs.* in healthy controls^50^. Austhof et al. reported the incidence of post-infection IBS to be 20.4% in a smaller study of 49 participants who completed a survey at six months, with half of these meeting the Rome IV diagnostic criteria for IBS^54^.

## Discussion

In this systematic review of 28 studies and 134,012 patients, we identified a relatively low (median = 6.25%) and variable (IQR = 10.2%; range 1.2–11.4%) prevalence of persistent gastrointestinal symptoms following SARS-CoV-2 infection. Prevalence estimates for persistent symptoms of any duration ranged from 0.2% to 24.1% for six studies judged to be at a low risk of bias, with a median follow-up time of 13 weeks. Functional gastrointestinal disorders, such as IBS and dyspepsia, were also observed after SARS-CoV-2 infection.

We found extensive inter-study heterogeneity, as expected when synthesising data from multiple observational studies across a variety of settings in changing circumstances. The studies with the highest methodological quality indicated that substantial numbers of individuals may be susceptible to persistent post-COVID gastrointestinal symptoms. We identified a higher pooled prevalence of diarrhoea, abdominal pain, nausea and vomiting in adults previously exposed to SARS-CoV-2 compared to controls. However, we do not report a reliable pooled effect estimate due to inter-study heterogeneity. Given the rapid spread of SARS-CoV-2 globally and recent UK estimates of post-COVID syndrome prevalence of 3.3%, a substantial proportion of those with chronic gastrointestinal complaints in the general population may be attributable to SARS-CoV-2^3, 58^.

Our review aimed to estimate the prevalence of persistent post-COVID gastrointestinal symptoms, but we faced several methodological challenges. Post-COVID syndrome is a dynamic condition that relapses and remits over time, so point prevalence estimates may not capture its true burden^59^. Half of the studies included were based on hospital cohorts of patients suffering more severe disease, which may overestimate symptom prevalence due to ascertainment bias. Our review supports the observations that post-COVID syndrome is more common in patients with a history of severe COVID-19^37, 47^. Hospitalised patients would generally be more likely to suffer comorbidities, including presentations with gastrointestinal symptoms^60^. Finally, four cross-sectional studies included in our review may be at a higher risk of recall bias, given the nature of recalling details of COVID-19 infection and providing a clinical history of gastrointestinal complaints retrospectively^30, 31, 55, 56^.

The prevalence of post-COVID syndrome may vary depending on the SARS-CoV-2 variant and the healthcare seeking behaviour of the population. COVID-19 illness severity and hospitalisation rates are known to differ between the original wild-type SARS-CoV-2 and later variants^61, 62^. As such, SARS-CoV-2 variant epochs may account for much of the variation in symptom prevalence over time and place that we found in this review. Healthcare seeking behaviour was also known to change throughout the COVID-19 pandemic; healthcare avoidance may result in an underestimation of the true frequency of gastrointestinal events in a population, particularly for studies requiring an in-person clinical review^63^.

Of the studies reporting incidence of gastrointestinal illness post-COVID-19, one found that, in children attending the emergency department with functional abdominal pain, those with historical SARS-CoV-2 infection were significantly more likely to be diagnosed with IBS than abdominal migraine, compared to pre-pandemic controls^44^. This finding is in keeping with the hypothesis that SARS-CoV-2 infection may trigger post-infection functional gastrointestinal symptoms, whereas the aetiology of abdominal migraine is more likely to be non-infectious^64^. Similarly, two studies reported a higher incidence of IBS between three and six months after COVID-19, however one study in our review did not find any significant difference.

In contrast, a cohort study of children assessing self-reported symptoms, suggested previous SARS-CoV-2 infection protects against persistent diarrhoea and nausea one month after subsequent COVID-19 diagnosis. Although younger child age has been shown to be protective for COVID-19 mortality, this study did not consider that SARS-CoV-2 positive children and their families were likely to be isolated from other causes of infectious diarrhoea during the study period^52, 65^.

It was not possible to estimate the impact of prior SARS-CoV-2 infection on other infectious or autoimmune mediated gastrointestinal illnesses based on the data presented in our review. Although limited, current research suggests that rates of *Clostridium difficile* infection are no different between SARS-CoV-2 exposed and unexposed patients^66^. However, no studies in our review reported the incidence of acute gastrointestinal infection following COVID-19, despite gut microbiome dysbiosis and prolonged inflammatory response being established sequelae. It is likely that non-pharmaceutical interventions (NPIs) intended to reduce COVID-19 transmission have also significantly reduced the transmission and case rates of enteric infections^67^. Therefore, if there is any excess risk and burden of enteric disease associated with SARS-CoV-2 infection, this would not be detected in these studies and may warrant further longitudinal follow-up and investigation. No studies reported the incidence of autoimmune mediated gastrointestinal illness, such as coeliac disease or inflammatory bowel disease, despite systemic viral and enteric infection being a proposed aetiology for both^68, 69^.

Most studies did not control for antibiotic exposure during acute SARS-CoV-2 infection, despite up to three-quarters of COVID-19 patients receiving antibiotics early in the pandemic^70^. One study included in our review found that individuals treated with antibiotics during acute infection were more likely to have persistent diarrhoea than those who did not; another identified antibiotic exposure as the strongest predictor for post-COVID gastrointestinal sequalae^47, 49^. Interestingly, Ghoshal et al reported that all patients with persistent dysphagia received either oral or intravenous antibiotics during acute COVID-19^50^. None of these studies, however, reported the incidence of bacterial co-infection in patients who received antibiotics.

Although not included in our initial extraction criteria, post-hoc analysis identified only one study reporting the incidence of low mood among patients with persistent post-COVID gastrointestinal symptoms, or vice-versa. This study did not find any significant associations between persistent gastrointestinal symptoms and symptoms of low mood, anxiety and sleep disturbance^49^. This is surprising considering that a higher prevalence of mental health conditions has been reported among those with functional gastrointestinal disorders during COVID-19 lockdowns, and emerging evidence of mechanisms linking depression and gut health^71, 72^.

## Conclusion

Our review found a generally low (median = 6.25%, IQR = 10.2%; range 1.2 – 11.4%) prevalence of persistent gastrointestinal symptoms up to twelve months after COVID-19 recovery. However, most studies lacked comparator groups, so we could not determine whether this differs from background rates of gastrointestinal illness in the general population. We did identify – albeit from limited data – that individuals previously exposed to SARS-CoV-2 may be more likely to develop IBS and dyspepsia than the general population. Significant heterogeneity between studies overall prevented us from providing reliable pooled estimates of long-lasting gastrointestinal consequences of SARS-CoV-2 infection. Ideally, COVID-19 studies would have included prospective observation of SARS-CoV-2 infected participants for the development of gastrointestinal complaints, accounting for vaccination, antimicrobial use, variant epochs and public health interventions. Given the established links between gut dysbiosis and a wide range of viral infections, new studies should also monitor excess risk of enteric infections after removal of COVID-related NPIs, and future pandemic preparedness would do well to include proactive surveillance of gastrointestinal infections.

## Data Availability

All data produced in the present work are contained in the manuscript

## Author Contributions

Conceptualisation, DH and IB; study design and methodology, DH, IB, NMV and MJH; literature and reference search, NMV and MJH; writing-preparation of the original draft, MJH; analysis, quality assessment, analysis, tables and figure preparation, LB, NMV and MJH; supervision, DH, DC, IB, AJE, LB. All authors read and approved the final manuscript.

## Data availability statement

All data produced in the present work are contained in the manuscript.

## References

1 Weekly epidemiological update on COVID-19 - 19 January 2023. Available at https://www.who.int/publications/m/item/weekly-epidemiological-update-on-covid-19---19-january-2023. Accessed April 11, 2023, n.d.

2 National Institute for Health and Care Excellence. Overview | COVID-19 rapid guideline: managing the long-term effects of COVID-19 | Guidance | NICE. Available at https://www.nice.org.uk/guidance/NG188. Accessed April 11, 2023, 2020.

3 Prevalence of ongoing symptoms following coronavirus (COVID-19) infection in the UK-Office for National Statistics. Available at https://www.ons.gov.uk/peoplepopulationandcommunity/healthandsocialcare/conditionsanddiseases/bulletins/prevalenceofongoingsymptomsfollowingcoronaviruscovid19infectionintheuk/5january2023. Accessed April 11, 2023, n.d.

4 Mair Manish, Singhavi Hitesh, Pai Ameya, Singhavi Jinesh, Gandhi Prachi, Conboy Peter, et al. A Meta-Analysis of 67 Studies with Presenting Symptoms and Laboratory Tests of COVID-19 Patients. The Laryngoscope 2021;131(6):1254–65. Doi: 10.1002/lary.29207.

5 Elshazli Rami M., Kline Adam, Elgaml Abdelaziz, Aboutaleb Mohamed H., Salim Mohamed M., Omar Mahmoud, et al. Gastroenterology manifestations and COVID-19 outcomes: A meta-analysis of 25,252 cohorts among the first and second waves. J Med Virol 2021;93(5):2740–68. Doi: 10.1002/jmv.26836.

6 Graham Gemma, Taegtmeyer Miriam, Lewis Joseph, Subramanian Sreedhar. Gastrointestinal symptoms involvement in hospitalised COVID-19 patients in Liverpool, UK: a descriptive cross-sectional, single-centre study. Clin Med (Lond) 2021;21(Suppl 2):23–4. Doi: 10.7861/clinmed.21-2-s23.

7 Aiyegbusi Olalekan Lee, Hughes Sarah E., Turner Grace, Rivera Samantha Cruz, McMullan Christel, Chandan Joht Singh, et al. Symptoms, complications and management of long COVID: a review. J R Soc Med 2021;114(9):428–42. Doi: 10.1177/01410768211032850.

8 Taquet Maxime, Dercon Quentin, Luciano Sierra, Geddes John R., Husain Masud, Harrison Paul J. Incidence, co-occurrence, and evolution of long-COVID features: A 6-month retrospective cohort study of 273,618 survivors of COVID-19. PLoS Med 2021;18(9):e1003773. Doi: 10.1371/journal.pmed.1003773.

9 Chopra Neha, Chowdhury Mohit, Singh Anupam K., Ma Khan, Kumar Arvind, Ranjan Piyush, et al. Clinical predictors of long COVID-19 and phenotypes of mild COVID-19 at a tertiary care centre in India. Drug Discov Ther 2021;15(3):156–61. Doi: 10.5582/ddt.2021.01014.

10 Meringer Hadar, Mehandru Saurabh. Gastrointestinal post-acute COVID-19 syndrome. Nat Rev Gastroenterol Hepatol 2022;19(6):345–6. Doi: 10.1038/s41575-022-00611-z.

11 Zou Xin, Chen Ke, Zou Jiawei, Han Peiyi, Hao Jie, Han Zeguang. Single-cell RNA-seq data analysis on the receptor ACE2 expression reveals the potential risk of different human organs vulnerable to 2019-nCoV infection. Front Med 2020;14(2):185–92. Doi: 10.1007/s11684-020-0754-0.

12 Scialo Filippo, Daniele Aurora, Amato Felice, Pastore Lucio, Matera Maria Gabriella, Cazzola Mario, et al. ACE2: The Major Cell Entry Receptor for SARS-CoV-2. Lung 2020;198(6):867–77. Doi: 10.1007/s00408-020-00408-4.

13 Hashimoto Tatsuo, Perlot Thomas, Rehman Ateequr, Trichereau Jean, Ishiguro Hiroaki, Paolino Magdalena, et al. ACE2 links amino acid malnutrition to microbial ecology and intestinal inflammation. Nature 2012;487(7408):477–81. Doi: 10.1038/nature11228.

14 Gu Silan, Chen Yanfei, Wu Zhengjie, Chen Yunbo, Gao Hainv, Lv Longxian, et al. Alterations of the Gut Microbiota in Patients With Coronavirus Disease 2019 or H1N1 Influenza. Clin Infect Dis 2020;71(10):2669–78. Doi: 10.1093/cid/ciaa709.

15 Zhou Yaya, Shi Xing, Fu Wei, Xiang Fei, He Xinliang, Yang Bohan, et al. Gut Microbiota Dysbiosis Correlates with Abnormal Immune Response in Moderate COVID-19 Patients with Fever. J Inflamm Res 2021;14:2619–31. Doi: 10.2147/JIR.S311518.

16 Hussain Ikram, Cher Gabriel Liu Yuan, Abid Muhammad Abbas, Abid Muhammad Bilal. Role of Gut Microbiome in COVID-19: An Insight Into Pathogenesis and Therapeutic Potential. Front Immunol 2021;12:765965. Doi: 10.3389/fimmu.2021.765965.

17 Zuo Tao, Zhang Fen, Lui Grace C. Y., Yeoh Yun Kit, Li Amy Y. L., Zhan Hui, et al. Alterations in Gut Microbiota of Patients With COVID-19 During Time of Hospitalization. Gastroenterology 2020;159(3):944–955.e8. Doi: 10.1053/j.gastro.2020.05.048.

18 Phetsouphanh Chansavath, Darley David R., Wilson Daniel B., Howe Annett, Munier C. Mee Ling, Patel Sheila K., et al. Immunological dysfunction persists for 8 months following initial mild-to-moderate SARS-CoV-2 infection. Nat Immunol 2022;23(2):210–6. Doi: 10.1038/s41590-021-01113-x.

19 Kumar Ashutosh, Kumari Chiman, Faiq Muneeb A., Pareek Vikas, Narayan Ravi K. SARS-CoV-2 Infectivity Vis-A-Vis Human Gut and Mesentery: Pathogenic Implications for COVID-19. SSRN Journal 2020. Doi: 10.2139/ssrn.3636978.

20 Lee Sunhee, Yoon Gun Young, Myoung Jinjong, Kim Seong-Jun, Ahn Dae-Gyun. Robust and persistent SARS-CoV-2 infection in the human intestinal brush border expressing cells. Emerg Microbes Infect 2020;9(1):2169–79. Doi: 10.1080/22221751.2020.1827985.

21 Pretorius Etheresia, Venter Chantelle, Laubscher Gert Jacobus, Kotze Maritha J., Oladejo Sunday O., Watson Liam R., et al. Prevalence of symptoms, comorbidities, fibrin amyloid microclots and platelet pathology in individuals with Long COVID/Post-Acute Sequelae of COVID-19 (PASC). Cardiovasc Diabetol 2022;21(1):148. Doi: 10.1186/s12933-022-01579-5.

22 Kell Douglas B., Laubscher Gert Jacobus, Pretorius Etheresia. A central role for amyloid fibrin microclots in long COVID/PASC: origins and therapeutic implications. Biochem J 2022;479(4):537–59. Doi: 10.1042/BCJ20220016.

23 Fikree Asma, Byrne Peter. Management of functional gastrointestinal disorders. Clin Med (Lond) 2021;21(1):44–52. Doi: 10.7861/clinmed.2020-0980.

24 Ballou Sarah, Bedell Alyse, Keefer Laurie. Psychosocial impact of irritable bowel syndrome: A brief review. World J Gastrointest Pathophysiol 2015;6(4):120–3. Doi: 10.4291/wjgp.v6.i4.120.

25 Fancourt Daisy, Steptoe Andrew, Bu Feifei. Long-term psychological consequences of long Covid: a propensity score matching analysis comparing trajectories of depression and anxiety symptoms before and after contracting long Covid vs short Covid 2022:2022.04.01.22273305. Doi: 10.1101/2022.04.01.22273305.

26 Migliavaca Celina Borges, Stein Cinara, Colpani Verônica, Munn Zachary, Falavigna Maicon, Prevalence Estimates Reviews – Systematic Review Methodology Group (PERSyst). Quality assessment of prevalence studies: a systematic review. J Clin Epidemiol 2020;127:59–68. Doi: 10.1016/j.jclinepi.2020.06.039.

27 Halverson Colleen C., Bailey Catherine, Ennis Joyce Arlene, Cox E. Elaine. Nursing surveillance of respiratory adverse events among hospitalized adults: A systematic review to guide evidence-based practice. Worldviews Evid Based Nurs 2022;19(4):260–6. Doi: 10.1111/wvn.12581.

28 Cheung Teris, Cheng Calvin Pak Wing, Fong Tommy Kwan Hin, Sharew Nigussie Tadesse, Anders Robert L., Xiang Yu Tao, et al. Psychological impact on healthcare workers, general population and affected individuals of SARS and COVID-19: A systematic review and meta-analysis. Front Public Health 2022;10:1004558. Doi: 10.3389/fpubh.2022.1004558.

29 Balduzzi Sara, Rücker Gerta, Schwarzer Guido. How to perform a meta-analysis with R: a practical tutorial. Evid Based Ment Health 2019;22(4):153–60. Doi: 10.1136/ebmental-2019-300117.

30 Islam Md. Saiful, Ferdous Most. Zannatul, Islam Ummay Soumayia, Mosaddek Abu Syed Md., Potenza Marc N., Pardhan Shahina. Treatment, Persistent Symptoms, and Depression in People Infected with COVID-19 in Bangladesh. Int J Environ Res Public Health 2021;18(4):1453. Doi: 10.3390/ijerph18041453.

31 Khodeir Mostafa M., Shabana Hassan A., Rasheed Zafar, Alkhamiss Abdullah S., Khodeir Mohamed, Alkhowailed Mohammad S., et al. COVID-19: Post-recovery long-term symptoms among patients in Saudi Arabia. PLoS One 2021;16(12):e0260259. Doi: 10.1371/journal.pone.0260259.

32 Qamar Mohammad Aadil, Martins Russell Seth, Dhillon Rubaid Azhar, Tharwani Areeba, Irfan Omar, Suriya Qosain Fatima, et al. Residual symptoms and the quality of life in individuals recovered from COVID-19 infection: A survey from Pakistan. Ann Med Surg (Lond) 2022;75:103361. Doi: 10.1016/j.amsu.2022.103361.

33 Vaillant Marie-France, Agier Lydiane, Martineau Caroline, Philipponneau Manon, Romand Dorothée, Masdoua Virginie, et al. Food intake and weight loss of surviving inpatients in the course of COVID-19 infection: A longitudinal study of the multicenter NutriCoviD30 cohort. Nutrition 2022;93:111433. Doi: 10.1016/j.nut.2021.111433.

34 Gérard Marine, Mahmutovic Meliha, Malgras Aurélie, Michot Niasha, Scheyer Nicolas, Jaussaud Roland, et al. Long-Term Evolution of Malnutrition and Loss of Muscle Strength after COVID-19: A Major and Neglected Component of Long COVID-19. Nutrients 2021;13(11):3964. Doi: 10.3390/nu13113964.

35 Damanti Sarah, Cilla Marta, Cilona Maria, Fici Aldo, Merolla Aurora, Pacioni Giacomo, et al. Prevalence of Long COVID-19 Symptoms After Hospital Discharge in Frail and Robust Patients. Front Med (Lausanne) 2022;9:834887. Doi: 10.3389/fmed.2022.834887.

36 Karaarslan Fatih, Demircioğlu Güneri Fulya, Kardeş Sinan. Postdischarge rheumatic and musculoskeletal symptoms following hospitalization for COVID-19: prospective follow-up by phone interviews. Rheumatol Int 2021;41(7):1263–71. Doi: 10.1007/s00296-021-04882-8.

37 Belkacemi Mohamed, Baouche Hayet, Gomis Sébastien, Lassalle Mathilde, Couchoud Cécile, the REIN registry. Long-lasting clinical symptoms 6 months after COVID-19 infection in the French national cohort of patients on dialysis. J Nephrol 2022;35(3):787–93. Doi: 10.1007/s40620-022-01295-z.

38 Wu Qiao, Ailshire Jennifer A., Crimmins Eileen M. Long COVID and symptom trajectory in a representative sample of Americans in the first year of the pandemic. Sci Rep 2022;12:11647. Doi: 10.1038/s41598-022-15727-0.

39 Faycal A., Ndoadoumgue A.L., Sellem B., Blanc C., Dudoit Y., Schneider L., et al. Prevalence and factors associated with symptom persistence: A prospective study of 429 mild COVID-19 outpatients. Infect Dis Now 2022;52(2):75–81. Doi: 10.1016/j.idnow.2021.11.003.

40 Robineau Olivier, Wiernik Emmanuel, Lemogne Cédric, de Lamballerie Xavier, Ninove Laetitia, Blanché Hélène, et al. Persistent symptoms after the first wave of COVID-19 in relation to SARS-CoV-2 serology and experience of acute symptoms: A nested survey in a population-based cohort. Lancet Reg Health Eur 2022;17:100363. Doi: 10.1016/j.lanepe.2022.100363.

41 Liang Limei, Yang Bohan, Jiang Nanchuan, Fu Wei, He Xinliang, Zhou Yaya, et al. Three-month Follow-up Study of Survivors of Coronavirus Disease 2019 after Discharge. J Korean Med Sci 2020;35(47):e418. Doi: 10.3346/jkms.2020.35.e418.

42 Xie Xiao-Ping, Sheng Li-Ping, Han Chao-Qun, Jin Yu, Bai Tao, Lin Rong, et al. Features of capsule endoscopy in COVID-19 patients with a six-month follow-up: A prospective observational study. J Med Virol 2022;94(1):246–52. Doi: 10.1002/jmv.27308.

43 Penner Justin, Abdel-Mannan Omar, Grant Karlie, Maillard Sue, Kucera Filip, Hassell Jane, et al. 6-month multidisciplinary follow-up and outcomes of patients with paediatric inflammatory multisystem syndrome (PIMS-TS) at a UK tertiary paediatric hospital: a retrospective cohort study. The Lancet Child & Adolescent Health 2021;5(7):473–82. Doi: 10.1016/S2352-4642(21)00138-3.

44 Stepan Mioara Desdemona, Cioboata Ramona, Vintilescu Ştefăniţa Bianca, Vasile Corina Maria, Osman Andrei, Ciolofan Mircea Sorin, et al. Pediatric Functional Abdominal Pain Disorders following COVID-19. Life (Basel) 2022;12(4):509. Doi: 10.3390/life12040509.

45 Comelli Agnese, Viero Giulia, Bettini Greta, Nobili Alessandro, Tettamanti Mauro, Galbussera Alessia Antonella, et al. Patient-Reported Symptoms and Sequelae 12 Months After COVID-19 in Hospitalized Adults: A Multicenter Long-Term Follow-Up Study. Front Med (Lausanne) 2022;9:834354. Doi: 10.3389/fmed.2022.834354.

46 Attauabi Mohamed, Dahlerup Jens Frederik, Poulsen Anja, Hansen Malte Rosager, Vester-Andersen Marianne Kajbæk, Eraslan Sule, et al. Outcomes and Long-Term Effects of COVID-19 in Patients with Inflammatory Bowel Diseases – A Danish Prospective Population-Based Cohort Study with Individual-Level Data. J Crohns Colitis 2021:jjab192. Doi: 10.1093/ecco-jcc/jjab192.

47 Liptak Peter, Duricek Martin, Rosolanka Robert, Ziacikova Ivana, Kocan Ivan, Uhrik Peter, et al. Gastrointestinal sequalae months after severe acute respiratory syndrome corona virus 2 infection: a prospective, observational study. European Journal of Gastroenterology & Hepatology 2022;34(9):925. Doi: 10.1097/MEG.0000000000002425.

48 Augustin Max, Schommers Philipp, Stecher Melanie, Dewald Felix, Gieselmann Lutz, Gruell Henning, et al. Post-COVID syndrome in non-hospitalised patients with COVID-19: a longitudinal prospective cohort study. Lancet Reg Health Eur 2021;6:100122. Doi: 10.1016/j.lanepe.2021.100122.

49 Noviello Daniele, Costantino Andrea, Muscatello Antonio, Bandera Alessandra, Consonni Dario, Vecchi Maurizio, et al. Functional gastrointestinal and somatoform symptoms five months after SARS-CoV-2 infection: A controlled cohort study. Neurogastroenterol Motil 2022;34(2):e14187. Doi: 10.1111/nmo.14187.

50 Ghoshal Uday C, Ghoshal Ujjala, Rahman M Masudur, Mathur Akash, Rai Sushmita, Akhter Mahfuza, et al. Post-infection functional gastrointestinal disorders following coronavirus disease-19: A case–control study. J Gastroenterol Hepatol 2022;37(3):489–98. Doi: 10.1111/jgh.15717.

51 Galván-Tejada Carlos E., Herrera-García Cintya Fabiola, Godina-González Susana, Villagrana-Bañuelos Karen E., Amaro Juan Daniel De Luna, Herrera-García Karla, et al. Persistence of COVID-19 Symptoms after Recovery in Mexican Population. Int J Environ Res Public Health 2020;17(24):9367. Doi: 10.3390/ijerph17249367.

52 Borch Luise, Holm Mette, Knudsen Maria, Ellermann-Eriksen Svend, Hagstroem Soeren. Long COVID symptoms and duration in SARS-CoV-2 positive children — a nationwide cohort study. Eur J Pediatr 2022;181(4):1597–607. Doi: 10.1007/s00431-021-04345-z.

53 Fernández-Plata Rosario, Higuera-Iglesias Anjarath-Lorena, Torres-Espíndola Luz María, Aquino-Gálvez Arnoldo, Velázquez Cruz Rafael, Camarena Ángel, et al. Risk of Pulmonary Fibrosis and Persistent Symptoms Post-COVID-19 in a Cohort of Outpatient Health Workers. Viruses 2022;14(9):1843. Doi: 10.3390/v14091843.

54 Austhof Erika, Bell Melanie L., Riddle Mark S., Catalfamo Collin, McFadden Caitlyn, Cooper Kerry, et al. Persisting gastrointestinal symptoms and post-infectious irritable bowel syndrome following SARS-CoV-2 infection: results from the Arizona CoVHORT. Epidemiol Infect 2022;150:e136. Doi: 10.1017/S0950268822001200.

55 Rao Guduru Venkat, Gella Vishwanath, Radhakrishna Madhuri, Kumar V Jagdeesh, Chatterjee Robin, Kulkarni Anand, et al. Post-COVID-19 symptoms are not uncommon among recovered patients-A cross-sectional online survey among the Indian populatio 2021. Doi: 10.1101/2021.07.15.21260234.

56 Fatima Ghizal, Bhatt Divyansh, Idrees Jaserah, Khalid Bushra, Mahdi Farzana. Elucidating Post-COVID-19 manifestations in India 2021. Doi: 10.1101/2021.07.06.21260115.

57 Koloski N. A., Jones M., Hammer J., von Wulffen M., Shah A., Hoelz H., et al. The Validity of a New Structured Assessment of Gastrointestinal Symptoms Scale (SAGIS) for Evaluating Symptoms in the Clinical Setting. Dig Dis Sci 2017;62(8):1913–22. Doi: 10.1007/s10620-017-4599-6.

58 Daines Luke, Zheng Bang, Pfeffer Paul, Hurst John R., Sheikh Aziz. A clinical review of long-COVID with a focus on the respiratory system. Curr Opin Pulm Med 2022;28(3):174–9. Doi: 10.1097/MCP.0000000000000863.

59 Davis Hannah E., Assaf Gina S., McCorkell Lisa, Wei Hannah, Low Ryan J., Re’em Yochai, et al. Characterizing long COVID in an international cohort: 7 months of symptoms and their impact. EClinicalMedicine 2021;38:101019. Doi: 10.1016/j.eclinm.2021.101019.

60 Wong Martin Chi-Sang, Huang Junjie, Wong Yuet-Yan, Wong Grace Lai-Hung, Yip Terry Cheuk-Fung, Chan Rachel Ngan-Yin, et al. Epidemiology, Symptomatology, and Risk Factors for Long COVID Symptoms: Population-Based, Multicenter Study. JMIR Public Health Surveill 2023;9:e42315. Doi: 10.2196/42315.

61 Lin Lixin, Liu Ying, Tang Xiujuan, He Daihai. The Disease Severity and Clinical Outcomes of the SARS-CoV-2 Variants of Concern. Front Public Health 2021;9:775224. Doi: 10.3389/fpubh.2021.775224.

62 Aleem Abdul, Akbar Samad Abdul Bari, Vaqar Sarosh. Emerging Variants of SARS-CoV-2 And Novel Therapeutics Against Coronavirus (COVID-19). StatPearls. Treasure Island (FL): StatPearls Publishing; 2023.

63 Splinter Marije J., Velek Premysl, Ikram M. Kamran, Kieboom Brenda C. T., Peeters Robin P., Bindels Patrick J. E., et al. Prevalence and determinants of healthcare avoidance during the COVID-19 pandemic: A population-based cross-sectional study. PLoS Med 2021;18(11):e1003854. Doi: 10.1371/journal.pmed.1003854.

64 Mani Jyoti, Madani Shailender. Pediatric abdominal migraine: current perspectives on a lesser known entity. Pediatric Health Med Ther 2018;9:47–58. Doi: 10.2147/PHMT.S127210.

65 Khera Nina, Santesmasses Didac, Kerepesi Csaba, Gladyshev Vadim N. COVID-19 mortality rate in children is U-shaped. Aging (Albany NY) 2021;13(16):19954–62. Doi: 10.18632/aging.203442.

66 Granata Guido, Petrosillo Nicola, Al Moghazi Samir, Caraffa Emanuela, Puro Vincenzo, Tillotson Glenn, et al. The burden of Clostridioides difficile infection in COVID-19 patients: A systematic review and meta-analysis. Anaerobe 2022;74:102484. Doi: 10.1016/j.anaerobe.2021.102484.

67 Love Nicola K., Elliot Alex J., Chalmers Rachel M., Douglas Amy, Gharbia Saheer, McCormick Jacquelyn, et al. Impact of the COVID-19 pandemic on gastrointestinal infection trends in England, February-July 2020. BMJ Open 2022;12(3):e050469. Doi: 10.1136/bmjopen-2021-050469.

68 Kahrs Christian R, Chuda Katerina, Tapia German, Stene Lars C, Mårild Karl, Rasmussen Trond, et al. Enterovirus as trigger of coeliac disease: nested case-control study within prospective birth cohort. BMJ 2019:l231. Doi: 10.1136/bmj.l231.

69 Castaño-Rodríguez Natalia, Kaakoush Nadeem O, Lee Way Seah, Mitchell Hazel M. Dual role of Helicobacter and Campylobacter species in IBD: a systematic review and meta-analysis. Gut 2017;66(2):235–49. Doi: 10.1136/gutjnl-2015-310545.

70 Langford Bradley J, So Miranda, Raybardhan Sumit, Leung Valerie, Soucy Jean-Paul R, Westwood Duncan, et al. Antibiotic prescribing in patients with COVID-19: rapid review and meta-analysis. Clin Microbiol Infect 2021;27(4):520–31. Doi: 10.1016/j.cmi.2020.12.018.

71 Alzahrani Mohammed A., Alshamrani Ali S., Ahmasani Ibrahim M., Alahmari Fahad S., Asiri Ali H., Alshehri Abdullah M., et al. Coronavirus disease 2019 pandemic stress and its effects on irritable bowel syndrome patients in Saudi Arabia. Medicine (Baltimore) 2020;99(51):e23711. Doi: 10.1097/MD.0000000000023711.

72 Peirce Jason M., Alviña Karina. The role of inflammation and the gut microbiome in depression and anxiety. Journal of Neuroscience Research 2019;97(10):1223–41. Doi: 10.1002/jnr.24476.

